# Using local and statewide Electronic Health Record data to evaluate the impact of telemedicine in Virginia

**DOI:** 10.64898/2026.01.08.26343531

**Authors:** Saurav Sengupta, Johanna Loomba, Andrea Zhou, Richard Holtzclaw, Jacqueline Gravitt, Myron Chang, Joshua Eassa, David Driscoll, Hannah O’Donnell, Shounak Chattopadhyay, Karen Rheuban, Donald E. Brown

## Abstract

**Objective:** To analyze the impact of telemedicine on emergency department (ED) utilization among University of Virginia (UVA) Health System patients, examining which patient characteristics predict reduced ED usage and whether telemedicine reduces ED utilization.

**Materials and Methods:** We used UVA Electronic Health Records and public datasets to establish clinical and contextual features including demographics, comorbidities, insurance status, and community characteristics. UVA patient data were linked to Virginia Health Information (VHI) data at the individual level, ensuring our utilization measure included ED encounters across all Virginia health systems. We evaluated: (1) patient characteristics associated with reduced ED usage following the first telemedicine encounter using an XGBoost model and (2) associations between telemedicine and ED usage using fixed effects modeling.

**Results:** Younger, healthier patients with high prior ED usage experienced the greatest reduction in ED visits following their first telemedicine visit. Telemedicine was significantly associated with reduced ED utilization across all observation windows (3, 6, and 12 months), with effects attenuating over longer windows.

**Discussion:** Data pipelines and models were designed to support rapid iteration on varying feature sets and sub-populations and to enable longitudinal model retraining and evaluation. These findings suggest telemedicine reduces ED utilization with significant reductions observed in specific sub-populations in our cohort.

**Conclusion:** Telemedicine engagement is associated with meaningful reductions in ED utilization, particularly among younger, healthier high-utilizer patients. Data science tools can help providers and policymakers optimize telemedicine delivery to benefit patients while reducing health system burden.

## INTRODUCTION

Limited access to care contributes to preventable ED use, which strains healthcare systems ^1^ and reflects sub-optimal patient management ^2^. The Virginia Rural Health Plan (2022–2026) highlights a critical disparity: rural communities have some of the lowest rates of primary care use but the highest rates of emergency department (ED) visits ^3^. Telemedicine has been proposed as a solution for addressing these health disparities between rural and urban populations ^4,5^. The National Committee for Quality Assurance (NCQA) Taskforce for Telehealth Policy (TTP) cites studies by several different health systems and insurance organizations. Such studies show that individuals with access to virtual care are notably less likely to seek care via the ED, effectively reducing care costs ^6^. Thus prior evidence suggests that enhancing access through telemedicine services is associated with reduced ED utilization. However, little has been done to build models that identify the patients that are most impacted by increased access or to establish a causal relationship between access to telemedicine and reduced ED utilization.

The UVA Center for Telehealth ^7^ has been instrumental in expanding telemedicine services statewide, particularly to patients in more remote regions of the Commonwealth of Virginia. Virginia Medicaid was an early adopter of telemedicine reimbursement in 2003, and with a legislative commercial plan mandate for coverage in 2010, health systems began to adopt telemedicine as a care delivery model. However, the broader policy changes implemented during the COVID-19 Public Health Emergency (PHE) included permanent mandates for Medicaid and commercial plan coverage of telemedicine services in Virginia, regardless of geographic or originating site location (including the home), and with the waivers implemented by Medicare, utilization of telemedicine services scaled exponentially statewide. The Medicare policy waivers have not yet been made permanent but rather have been maintained via a federal continuing resolution. As such, policy decisions surrounding Medicare coverage of telemedicine must continue to be informed by data on utilization and outcomes. We hypothesized that many factors both at the individual patient and regional level might elucidate who is most impacted by care provided via telemedicine. In order to strategically expand or target telemedicine services, and to inform policy decisions, our multidisciplinary team of informaticians, data scientists, and clinicians developed data pipelines and AI tools to model how UVA patient features predicted their Virginia ED utilization trends before and after telemedicine engagement.

Patient encounter data stored in the UVA Electronic Health Record (EHR) was used to capture telemedicine services as they expanded and evolved over time. Individual patient features such as their demographic characteristics, chronic comorbidities, and diagnoses most often associated with UVA telemedicine visits were also derived from the EHR data. We included Medicaid and Medicare coverage, which have been correlated with infrastructure related barriers to care in patients seen for non-urgent needs in the ED ^1^. Regional features were also included for each patient, such as rurality and community conditions, so that our models could account for individual health status while controlling for regional features that can also correlate with access to care ^8^.

In order to obtain ED visits from all hospital systems in the state, we leveraged Virginia Health Information (VHI) Exchange data linked to our UVA EHR data. Because each dataset covered different time periods (both as a whole, and in terms of individual patient observation periods), we carefully selected subsets of our cohort for each analysis to ensure only those with adequate longitudinal data in both datasets were included.

## MATERIALS AND METHODS

### Data Sources and Derived Features

#### University of Virginia (UVA) EHR Derived Features

We used data from the University of Virginia (UVA) Electronic Health Record (EHR) system to capture patient demographic, clinical, and telemedicine encounter features. Due to evolving workflows, billing, and coding practices over the years, as well as some variation in definition, accurately identifying telemedicine encounters in source data can be challenging. For this study, we chose to define telemedicine as any audio or video direct care encounter, which excluded events like asynchronous electronic communication with a clinician or remote patient monitoring. Through data exploration, chart review, and expert consults from operational teams, we established that any of the following were specific indicators of visits that met our definition:

1. visit in the Department of UVPC Telemedicine
2. visit with TELEHEALTH_MODE Code of Video, Telephone, Clinic to Clinic Video
3. visit that have smartphrases in the provider note containing the subphrases TELEMEDVIDEO or TELEPHCALL at any time
4. visit with an encounter type that contains “telemedicine”

These criteria were developed in close partnership with the UVA Karen Rheuban Center for Telehealth, UVA EHR analysts, and billing parties who were familiar with UVA telemedicine coding practices as they evolved.

Telemedicine billing codes were first introduced at UVA more than 25 years ago, but UVA integrated home as an eligible patient originating site following the PHE. Since then, identification of telemedicine visits has been simplified. Visits with mode codes of In-Person, eVisit, and Patient Not Present were excluded from this study as they did not accurately reflect audio or video direct care encounters. TELEMEDVIDEO and TELEPHCALL smartphrases populate the provider note with the attestation language that the provider obtained the patient’s consent for a telemedicine encounter. In-Person visits were occasionally converted last minute to virtual care without having the TELEHEALTH_MODE Code updated so we use the smartphrase criteria to catch these outliers where the mode had not been updated. While eVisits may sound as though they do qualify as tele-health, visits with this classification in the UVA EHR were predominantly asynchronous electronic communications with various members of the care team from scheduling staff to the clinicians. Other codes were considered, but not included in our algorithm due to lack of specificity or the fact that they did not add additional visits.

UVA health records are mapped to the Observational Medical Outcomes Partnership (OMOP) ^9^ Common Data Model (CDM). A telemedicine visit type was added to our UVA OMOP encounter data based on the above algorithm. OMOP data were extracted for all UVA patients with at least one telemedicine visit, resulting in a source dataset containing raw structured HER data for 163,247 patients spanning 2010 to the date of extract (January 31, 2025). Table 5 in the supplemental materials provides detailed summary statistics of these patients and shows a slight majority female population with broadly distributed age groups. Figure 1 shows that UVA telemedicine visits from our study period are largely primary care visits but include a variety of other specialties as well.

**Figure 1.**
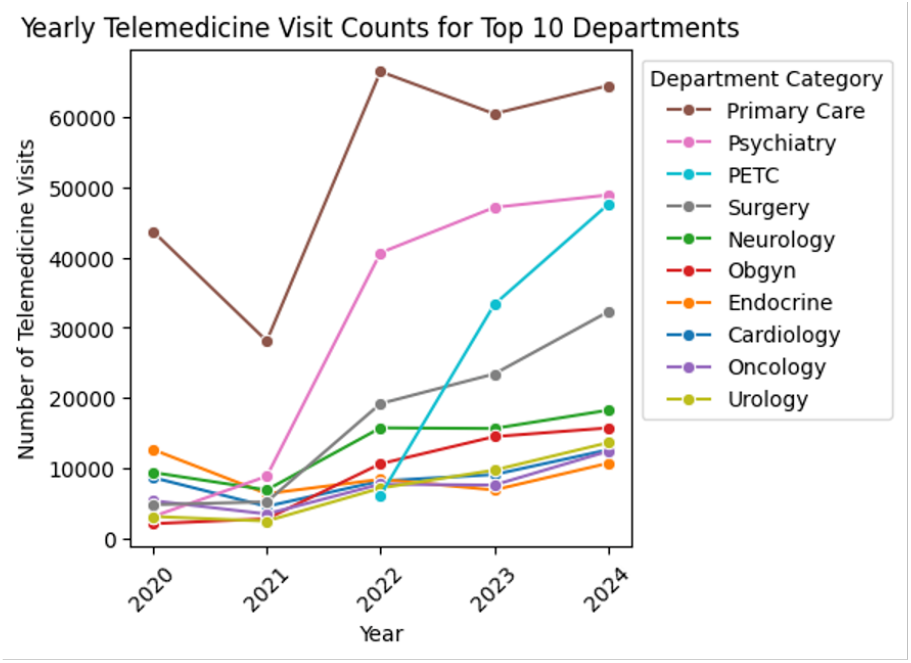
UVA Telemedicine Visits by Type and over time

The OMOP data for this cohort were also used to create our patient feature set. Because telemedicine has been shown to improve outcomes in patients with chronic disease ^10^, we calculated Charlson’s Comorbidity Index (CCI) ^11^ scores for our patients as captured over time. Due to the intermittent nature of data capture in the EHR, we created an accumulative score that included conditions in all future scores for the patient once recorded. The clinicians were also interested in finding population-specific clinical features so that we could control for the types of conditions that were most associated with telemedicine care at our local institution. Figure 2 shows the results of an exploration of the raw OMOP data.

**Figure 2.**
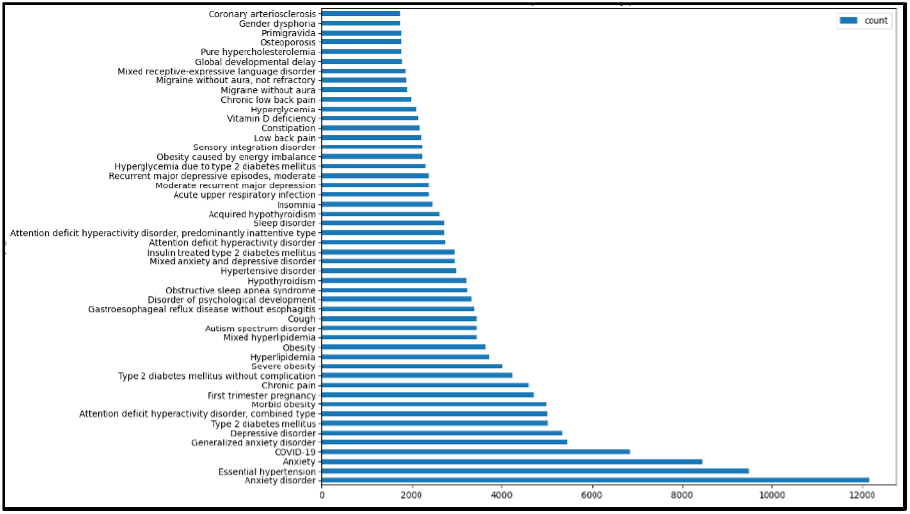
Top 50 conditions (y-axis) recorded during telemedicine visit by unique patient count (x-axis)

Based on this exploration, the team developed concept sets for: Chronic Pain, Peripheral Vascular Disease or Hyperlipidemia, Seizure Disorders, Sleep Disorders, Chronic Kidney Disease, Mood Disorders, Neurodevelopmental Disorders, and Diabetes, Pre-Diabetes, or Obesity. The UVA EHR also provided other patient features such as Medicare or Medicaid enrollment, age, sex, and zip code.

#### Virginia Health Information (VHI) Emergency Department Visits data

Virginia Health Information (VHI) stewards a Health Information Exchange (HIE) that has Emergency Department visits data in all hospital systems across Virginia for all UVA patients starting in August 2020. With VHI and UVA Institutional Review Board permission, we were able to link cross state ED visits to our University of Virginia (UVA) Health System patient data. Because VHI data provide information on ED utilization outside of UVA and because not all patients in our telemedicine cohort exclusively utilize the UVA Health Emergency Department, mapping statewide ED visit data to UVA Health patients was essential. Figure 3 show the magnitude of enhancement provided by the VHI data linkage.

**Figure 3.**
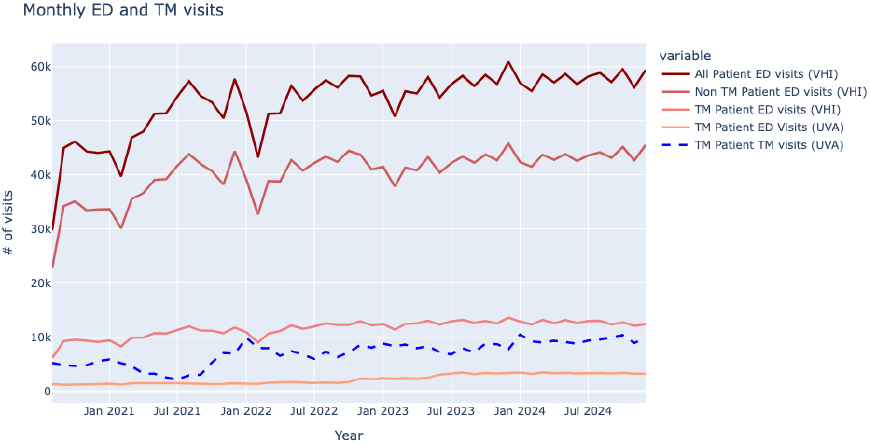
Monthly ED and TM visits, showing: 1. All UVA patients’ VHI ED visits, 2. Non-telemedicine UVA patients’ VHI ED visits, 3. UVA telemedicine patients’ ED visits from VHI, 4. Telemedicine patients with ED data only from UVA and 5. Telemedicine patients with Telemedicine visits (in blue). We can see how VHI data linkage increases ED visit counts compared to only using UVA data.

#### Regional Data

Each patient’s zip code was used to link public data recorded at the regional level. Because of the above cited associations between rurality and healthcare access in Virginia, we utilize Rural Urban Continuum Codes ^12^ to classify if patients are from a rural or urban region. A patient is given a value of 1 for the “rural” flag if they reside in an area with an urban population of 5,000 or less, adjacent or non-adjacent to a metro area. Community Condition Scores were also linked from the University of Wisconsin’s County Health Rankings ^13,14^. Community Condition Scores reflect social and economic factors, physical environment and health infrastructure in the region where the patient lives.

### Analysis 1: Which patients are helped most by telemedicine?

#### Cohort inclusion criteria

We defined inclusion criteria that were as broad as possible while ensuring that our outcome variable was meaningful.

We use the following inclusion criteria for this modeling:

1. All ages are included in this UVA patient analysis.
2. Adequate observation (temporal coverage of the patient sample):
  a. Only patients with at least one UVA encounter (any type) 28 days preand postfirst telemedicine visit
  b. Telemedicine index (TM index) should be >= 3 months after the VHI ED data start (effectively the index must be Nov 2020 or later)
3. Virginia (VHI) Emergency Department Encounters:
  a. Because we are looking at REDUCTION in ED use patterns for first analysis, we only included patients with at least one VHI ED visit

This results in a total patient cohort of 61,249 patients. Table 5 in supplemental materials shows the summary statistics of this cohort and their feature set.

#### Outcome variable

In order to identify the impact of our feature set on ED utilization, we define an outcome based on change of ED rates. Each patient has an average number of ED visits in their pre and post TM index observation periods. Our outcome variable y is defined as 1 when the visit rate drops in the post TM index period as compared to the pre-TM index period, and 0 otherwise. This provides a binary outcome, where reduced ED visit rates are the positive outcome, while increased ED visit rates are the negative outcome. The visits data for a sample patient is provided in Figure 4.

**Figure 4.**
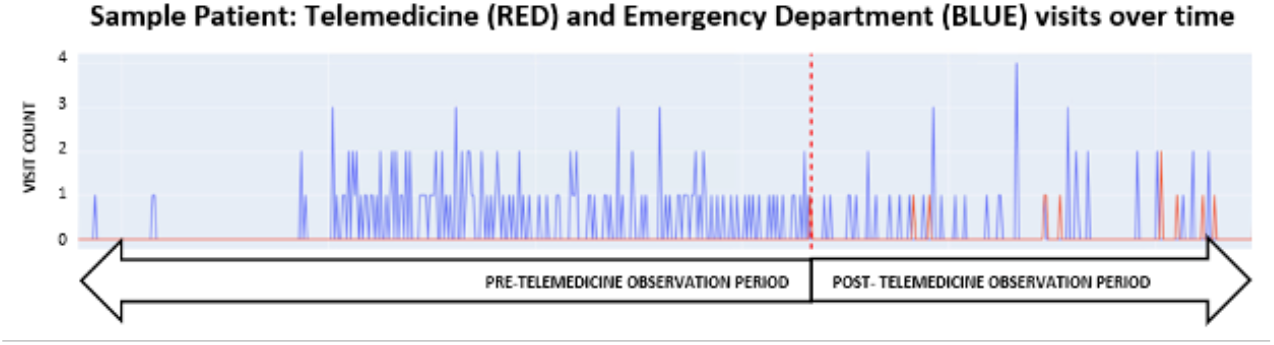
ED visit counts in blue and telemedicine visit counts in red for a sample patient from our cohort. Vertical dotted line denotes first UVA telemedicine visit for this patient. For this sample patient, their average rate is lower post-telemedicine which results in an outcome of 1.

#### Input features

The following features were used for this first analysis (refer to “Data Source and Derived Features” section):

1. Patient sex
2. Patient age at the time of TM index
3. Conditions associated with UVA telemedicine visits
4. Accumulative Charlson Comorbidity Index
5. Medicare and Medicaid flags
6. Time based features:
  a. Pre TM index ED visit rate (average since start of VHI ED data)
  b. Pre TM index observation period (time from VHI ED data start to TM index)
  c. Pre TM index ED visit rate since first ED visit (average time between ED visits prior to the TM index)
7. Community Condition Score (from County Health Rankings)
8. Rural flag

#### Model Design

We utilized an XGBoost ^15^ to build a classification model using the previously defined input features and the outcome variable. XGBoost was selected as it is robust to multicollinearity between features which we recognize as a strong possibility in this feature set. We then utilize Shapley Additive Explanations (SHAP) ^16^ to interpret model results to see which input features resulted in reduced ED visits. We divide our dataset into train and test sets in a 90%-10% split. We use 5-fold cross validation for our training data and evaluate the best fit model on our test data.

### Analysis 2: Does Telemedicine Impact ED Utilization?

As seen in Figure 5, there appears to be less of an upward trend in ED visits in our UVA patients who have engaged in telemedicine compared to those who have no record of a UVA telemedicine visit. We are therefore interested in modeling the association between the onset of telemedicine care and change in ED utilization. For this, we used a fixed effects model where we conditioned by patient ID to control for the characteristics of individuals (including both time invariant features and features that may change over time such as comorbidities). We looked at a fixed period of time from and before the start of telemedicine intervention (eg. 3 months before and after the first telemedicine visit) and captured the ED visit rates in the preand postintervention periods along with covariates mentioned in Analysis 1 above to feed into the model.

**Figure 5.**
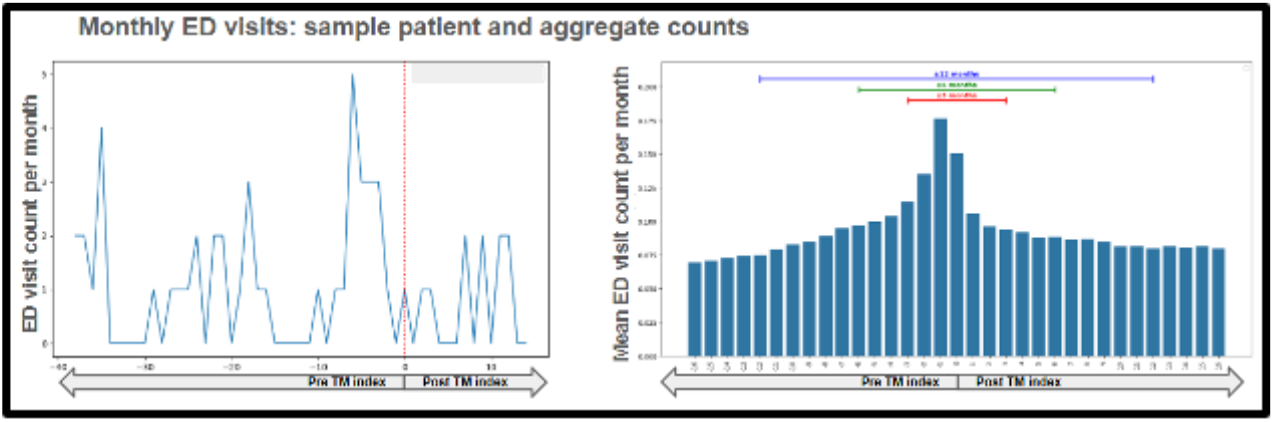
The chart on the left provides an example of ED visits in a month pre- and post-first telemedicine for a single patient. The chart on the right aggregates the monthly ED visit data for all patients in the cohort and aligns the counts around their TM index events. Intervals of 3, 6, and 12 months around the TM index are used for the below analyses.

#### Cohort inclusion criteria

To build this fixed effects model, we limited our cohort to patients with at least a year of potential ED observation (pre and post TM index) in the state-wide data. We use the following inclusion criteria for this modeling:

1. All ages are included in this UVA patient analysis.
2. TM index >= 365 days post start of VHI data and TM index <= 365 days prior to end of VHI data. This ensures we have sufficient observational data to compare pre and post intervention ED utilization

Note that unlike in the prior model eligibility, this cohort includes patients even if they had 0 visits in the pre-intervention period.

#### Outcome variable

We constructed a dataset by generating observation periods anchored to each patient’s telemedicine index date, creating balanced preand post-intervention windows of equal length (3, 6, or 12 months). For each patient-window combination, we calculated the ED visit rate by counting all ED encounters occurring within that period and dividing by the duration the patient was observed for in each window. The outcome variable captures visit frequency, allowing us to model within-patient changes in ED utilization patterns over time. Each patient contributes two observations to the dataset, one in the preintervention and one in the post-intervention, with a binary post-intervention indicator distinguishing the periods before versus after telemedicine initiation.

#### Input features

We calculated time-varying covariates at pre- and post-intervention, including the cumulative Charlson Comorbidity Index score, and comorbidities described in Analysis 1 based on diagnoses recorded up to that point in time. This structure enables the fixed effects model to identify treatment effects from within-patient temporal variation while controlling for patient-specific baseline characteristics that remain constant across the observation period.

#### Model Design

We employed a patient-level fixed effects panel regression model to estimate the causal effect of telemedicine intervention on emergency department (ED) visit rates across three observation windows (3, 6, and 12 months pre- and post-intervention). The fixed effects specification controls for all time-invariant patient characteristics—including baseline health status, demographics, and healthcare-seeking propensities—that could confound the relationship between telemedicine adoption and ED utilization, while the binary post-intervention indicator captures the within-patient change in ED visit rates following telemedicine initiation. We include time-varying covariates such as Charlson Comorbidity Index and others to account for temporal trends and evolving patient complexity. By comparing each patient to themselves before and after telemedicine exposure, we effectively control for unobserved patient-specific factors that might drive both telemedicine adoption and ED use, providing a more credible estimate of the intervention’s causal impact than cross-sectional comparisons. The multiple observation windows allow us to assess both immediate effects and sustained behavioral changes in healthcare utilization patterns.

## RESULTS AND DISCUSSION

### Analysis 1: Which patients are helped most by telemedicine?

Our model achieves a F1 score of 0.77 on the held out test set. We utilize SHAP plots to interpret the model (Figure 6).

**Figure 6.**
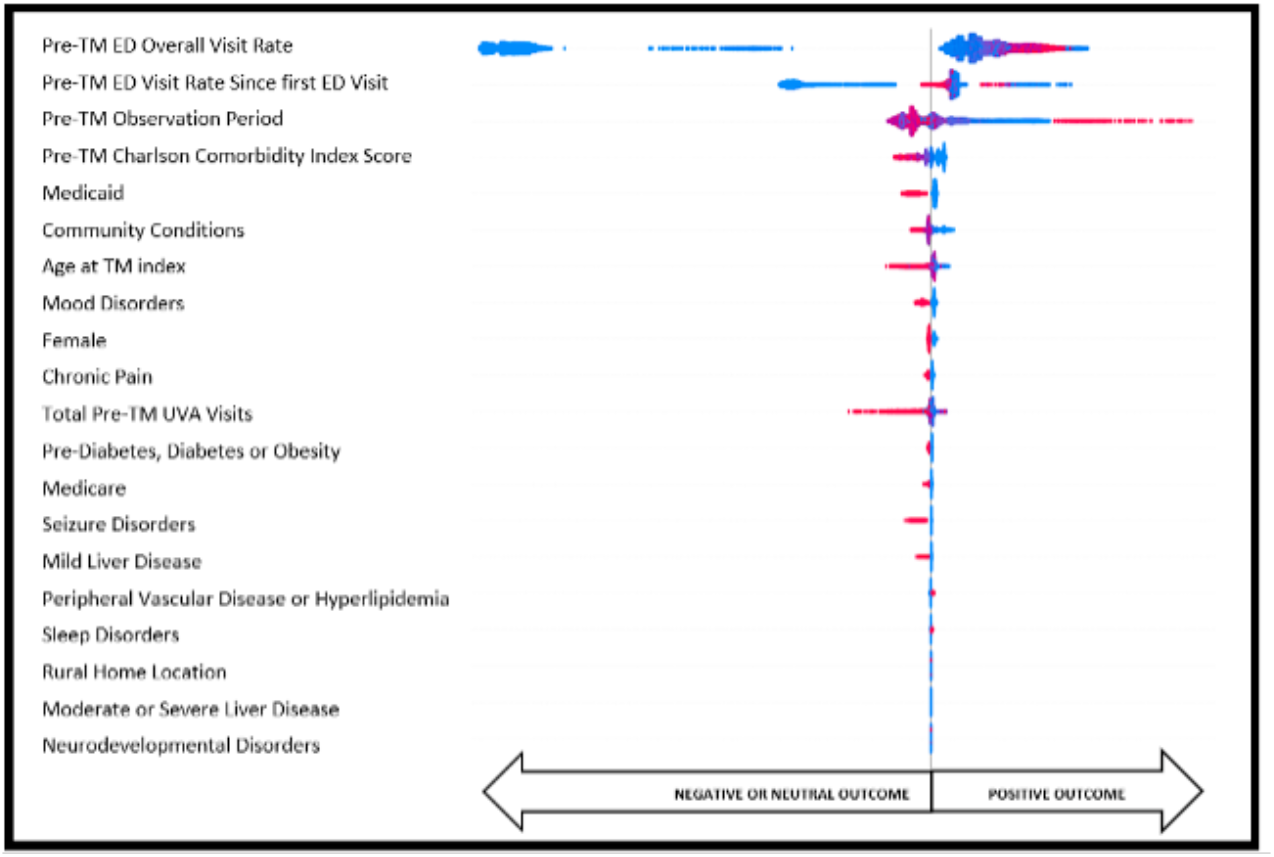
SHAP plots for full cohort show feature importance based on the XGBoost model after 10x cross validation. All features represent facts gathered prior to and up through the day of the first telemedicine encounter.

Each feature in the SHAP plot is associated with its impact on the outcome of “reduced ED rates post telemedicine” (y-axis). The blue points reflect samples where the value of the feature was low or zero (feature not present) and the red points represent samples where the value of the feature was high or one (feature present). Sample points to the right of midline were predicted by the model to have reduced ED visits post TM index. Thus, we see that the model largely associates positive outcomes (reduction in ED rates) with patients having the following features:

1. Lower ED visit rates prior to TM index, but greater prior engagement with the health system
2. “Healthier” patients with fewer chronic comorbidities (lower CCI score) and fewer telemedicine re-0.21 lated medical conditions (except for sleep disorders, which the model reports as being somewhat predictive of reduced ED visits)
3. Non-Medicaid
4. Lower Community Condition scores (better social, economic, physical environment and health infrastructure features in their locality)
5. Younger (lower age)

Note that the rurality of a patient’s home address is not a significant predictor of improved outcomes when we look at this broad patient sample that covers a large UVA catchment area and control for the other features. This suggests that telemedicine as a method to enhance access to care can impact patient outcomes across localities.

### Analysis 2: Does Telemedicine Impact ED Utilization?

Using a fixed effects panel regression model with the same cohort of 59,040 patients across three observation windows, we find that telemedicine adoption significantly reduces ED visit rates across all time windows applied, though the magnitude of effect substantially attenuates with longer windows. We present the results in Table 1.

**Table 1.** Fixed effects model results for post intervention indicator over all time windows for full cohort.

| Window | 3 month | 6 month | 12 month |
| --- | --- | --- | --- |
| Observations | 118080 | 118080 | 118080 |
| Patients | 59040 | 59040 | 59040 |
| Unadjusted Effect | -0.277 | -0.195 | -0.077 |
| Unadjusted SE | 0.018 | 0.012 | 0.009 |
| Adjusted Effect | -0.272 | -0.21 | -0.105 |
| Adjusted SE | 0.02 | 0.014 | 0.01 |
| Effect Difference | 0.005 | -0.015 | -0.028 |
| Percent Change | 1.80% | -7.70% | -36.40% |
| R<sup>2</sup> Overall | 0.693 | 0.743 | 0.783 |
| R<sup>2</sup> Within | 0.005 | 0.006 | 0.006 |
| RMSE | 2.134 | 1.5 | 1.121 |
| Mean Pre-Intervention Rate | 1.81 | 1.55 | 1.30 |
| Mean Post-Intervention Rate | 1.53 | 1.35 | 1.22 |

Mean pre-intervention ED visit rates vary depending on the time window applied with 1.81 visits/person-year in the 3-months prior window,1.55 in the 6-months prior, and 1.30 in12 months prior, reflecting the dynamic nature of the cohorts’ utilization patterns across different temporal frames. The adjusted treatment effects showed progressive attenuation: −0.272 visits/person-year at 3 months (15.0% reduction from mean pre index rates, p<0.001), −0.210 at 6 months (13.5% reduction, p<0.001), and −0.105 at 12 months (8.1% reduction, p<0.001), with effect sizes declining by 61% from the shortest to longest observation window. The increasing divergence between unadjusted and adjusted models over longer windows, from 1.8% at 3 months to 7.7% at 6 months and 36.4% at 12 months, indicates that time-varying confounders, particularly comorbidity burden measured by the Charlson Comorbidity Index and specific chronic conditions (seizures, chronic pain, sleep disorders, mood disorders), become progressively more important for longer observation periods as patient health status evolves. The patient-level fixed effects control for all time-invariant characteristics, with within-patient R^2^ remaining consistently low (0.005-0.006) across all models despite improving overall model fit (R^2^ Overall), confirming that most variation in ED utilization stems from stable individual differences rather than temporal changes. These findings suggest telemedicine has its strongest immediate impact on reducing ED visits, with the effect diminishing as behavioral patterns stabilize and underlying health trajectories increasingly assert themselves over the longer term.

Table 2 contains the coefficients for all time varying comorbid conditions added to the fixed effects model.

**Table 2.** Fixed effects model coefficients for post intervention indicator for 3/6/12 month windows for all patients in our cohort.

| Variable | 3 Month |  |  | 6 Month |  |  | 12 Month |  |  |
| --- | --- | --- | --- | --- | --- | --- | --- | --- | --- |
|  | Coef | SE | p-value | Coef | SE | p-value | Coef | SE | p-value |
| Post-Telemedicine Intervention Indicator | -0.272 | 0.020 | 0.000 | -0.210 | 0.014 | 0.000 | -0.105 | 0.010 | 0.000 |
| Neurological Disorders | 0.012 | 0.119 | 0.921 | 0.062 | 0.084 | 0.459 | 0.076 | 0.059 | 0.195 |
| Peripheral Vascular Disease or Hyperlipidemia | -0.169 | 0.087 | 0.051 | -0.056 | 0.062 | 0.363 | -0.022 | 0.044 | 0.612 |
| Seizure Disorders | 0.385 | 0.348 | 0.269 | 0.455 | 0.230 | 0.048 | 0.539 | 0.193 | 0.005 |
| Chronic Pain | 0.000 | 0.080 | 0.998 | 0.069 | 0.055 | 0.214 | 0.153 | 0.044 | 0.001 |
| Pre-Diabetes, Diabetes or Obesity | 0.008 | 0.078 | 0.918 | -0.006 | 0.057 | 0.923 | 0.007 | 0.048 | 0.880 |
| Sleep Disorders | 0.033 | 0.077 | 0.663 | 0.136 | 0.057 | 0.018 | 0.147 | 0.046 | 0.001 |
| Charlson's Comorbidity Score | 0.224 | 0.058 | 0.000 | 0.246 | 0.041 | 0.000 | 0.317 | 0.031 | 0.000 |
| Mood Disorders | 0.094 | 0.098 | 0.341 | 0.107 | 0.066 | 0.107 | 0.125 | 0.050 | 0.012 |
| Mild Liver Disease | -0.122 | 0.152 | 0.423 | -0.116 | 0.108 | 0.282 | -0.031 | 0.096 | 0.743 |
| Moderate/Severe Liver Disease | -0.590 | 0.365 | 0.106 | -0.226 | 0.254 | 0.372 | -0.023 | 0.183 | 0.902 |

The Charlson Comorbidity Index was the only covariate consistently significant across all observation windows, with coefficients increasing from 0.224 to 0.317 over longer periods, accounting for the growing importance of covariate adjustment (1.8% difference at 3 months versus 36.4% at 12 months). Seizure disorders, chronic pain, and sleep disorders exhibited time-dependent associations, becoming statistically significant only in longer observation windows, indicating de-layed or cumulative effects on ED utilization. Neurological conditions, peripheral vascular disease, and prediabetes/diabetes/obesity showed no significant associations in any window. Mood disorders emerge as a significant predictor only at 12 months (coefficient=0.125, p=0.012), contributing an additional 0.125 ED visits per year, while liver disease variables (mild and moderate/severe) show no significant associations at any time horizon despite their clinical relevance. The results indicate differential predictive patterns across time horizons: at 3 months, the intervention effect predominates with minimal comorbidity influence; at 6 months, intervention and comorbidity effects are balanced; at 12 months, comorbidity burden becomes the primary predictor while the intervention effect weakens substantially. These findings suggest that telemedicine reduces ED utilization in the immediate post-adoption period but does not produce sustained changes in long-term utilization patterns among patients with chronic conditions.

#### Sub-cohort analysis

To verify that the characteristics of patients most helped by telemedicine intervention identified by our first analysis we created a subcohort of patients with age at tele-health index of 18-44, no prior known comorbidities (cci pre index was 0) and high ED utilization (top quartile of pre telemedicine ED usage).

This analysis focuses on patients aged 18-44 with zero comorbidity burden (Pre-index CCI was zero) but high baseline ED utilization. We present the results for all windows for this sub-cohort in Tables 3 and 4.

**Table 3.** Fixed Effects Model Results Across All Time Windows - Young Healthy High ED Users.

| Window | Baseline ED Rate | Estimate | Std Error | P value | CI 2.5% | CI 97.5% | RMSE | R<sup>2</sup> Overall | R<sup>2</sup> Within |
| --- | --- | --- | --- | --- | --- | --- | --- | --- | --- |
| 3_month | 5.69 | -1.477 | 0.327 | 0.000 | -2.118 | -0.836 | 3.554 | 0.762 | 0.050 |
| 6_month | 5.57 | -1.637 | 0.262 | 0.000 | -2.152 | -1.122 | 2.910 | 0.810 | 0.097 |
| 12_month | 7.11 | -3.255 | 0.257 | 0.000 | -3.758 | -2.751 | 2.780 | 0.784 | 0.258 |

**Table 4.** Fixed Effects Model Results with Covariates Across Time Windows - Young Healthy High ED Users.

| Variable | 3 Month |  |  | 6 Month |  |  | 12 Month |  |  |
| --- | --- | --- | --- | --- | --- | --- | --- | --- | --- |
|  | Coef | SE | p-value | Coef | SE | p-value | Coef | SE | p-value |
| Post-Telemedicine Intervention Indicator | -1.477 | 0.327 | 0.000 | -1.637 | 0.262 | 0.000 | -3.255 | 0.257 | 0.000 |
| Neurological Disorders | 1.179 | 1.598 | 0.461 | -0.679 | 1.316 | 0.606 | -0.847 | 0.765 | 0.269 |
| Peripheral Vascular Disease or Hyperlipidemia | -1.220 | 2.012 | 0.545 | -0.613 | 1.066 | 0.565 | -1.776 | 1.054 | 0.092 |
| Seizure Disorders | 0.978 | 2.849 | 0.731 | 1.112 | 1.169 | 0.342 | -1.635 | 0.664 | 0.014 |
| Chronic Pain | 1.272 | 1.149 | 0.269 | 2.870 | 0.860 | 0.001 | 2.185 | 0.715 | 0.002 |
| Pre-Diabetes, Diabetes or Obesity | 0.072 | 1.570 | 0.963 | -1.016 | 1.403 | 0.469 | -0.780 | 1.211 | 0.520 |
| Sleep Disorders | 1.277 | 1.018 | 0.210 | 0.884 | 0.977 | 0.366 | 0.223 | 0.990 | 0.822 |
| Charlson's Comorbidity Score | -0.974 | 1.224 | 0.427 | 0.309 | 0.947 | 0.745 | 0.603 | 0.479 | 0.208 |
| Mood Disorders | -0.829 | 1.190 | 0.487 | 0.556 | 0.913 | 0.543 | 0.981 | 0.711 | 0.168 |
| Mild Liver Disease | -2.255 | 2.335 | 0.335 | -6.056 | 3.626 | 0.095 | -6.537 | 3.946 | 0.098 |
| Moderate/Severe Liver Disease | -8.290 | 2.414 | 0.001 | -9.622 | 3.359 | 0.004 | -4.203 | 3.587 | 0.242 |

Among 585 young adults (age 18-44) with zero comorbidity burden but high baseline ED utilization (5.57-7.11 visits/year across windows), telehealth adoption produced a pattern of increasing treatment effects over time, the opposite of temporal attenuation observed in our full cohort, with reductions growing from −1.477 visits/year at 3 months (p<0.001) to −1.637 at 6 months (p<0.001) and ultimately −3.255 at 12 months (p<0.001, representing a 45.8% reduction from baseline). Chronic pain emerged as the primary driver of residual ED utilization, becoming highly significant at 6 months (*β* = 2.870, p=0.001) and remaining significant at 12 months (*β* = 2.185, p=0.002), indicating that pain management represents a key barrier to complete ED substitution even among otherwise healthy telehealth adopters. The exceptional within-patient R^2^ of 0.258 at 12 months (compared to 0.005-0.009 in the overall population) demonstrates that telehealth intervention and covariates explain over a quarter of ED visit variation in this subgroup.

## CONCLUSION

In this analysis, we find that telemedicine is associated with a meaningful reduction in emergency department (ED) utilization among UVA patients across the state of Virginia. Using the first telemedicine encounter as an index intervention, our results demonstrate a statistically significant negative causal effect on ED visit rates. These findings support the assertion that telemedicine can serve as an effective alternative pathway for care that mitigates unnecessary ED use, with important implications for health system capacity, funding, and patient outcomes. The reduction in ED visits is not uniform across the population. The greatest impact is observed among younger, healthier patients who tend to use fewer healthcare services overall but have higher baseline rates of ED utilization. This suggests that telemedicine may be particularly effective for individuals whose acute or episodic care needs can be addressed outside of traditional emergency settings. Moreover, the stronger effect observed in this sub-cohort substantiates our analytic approach and highlights the value of stratified analyses in understanding differential impacts of telemedicine adoption. Importantly, this framework can be readily extended to examine telemedicine’s effects within specific sub-populations. By reapplying the model to patients in defined geographic regions or with particular clinical conditions, health systems can identify where telemedicine expansion is likely to yield the greatest benefit. The ability to control for evolving comorbidities over time further enables these methods to be used not only for identifying high-impact populations, but also for monitoring sustained effects as telemedicine programs mature.

Looking ahead, these findings provide a foundation for future work examining downstream outcomes, including reductions in hospitalizations and overall cost of care. More detailed analyses of cost savings and utilization patterns will be critical for informing public policy decisions related to telemedicine coverage and reimbursement. Together, these results underscore telemedicine’s potential as a scalable intervention for improving care delivery efficiency while reducing strain on emergency departments. We could employ lagged regression approaches with lag models or autoregressive specifications to explore dynamic temporal relationships between telehealth adoption timing and ED utilization patterns, testing whether treatment effects exhibit delayed onset, persistence across multiple periods, or feedback loops where prior ED visits predict subsequent telehealth engagement intensity. This helps us in distinguishing immediate behavioral substitution from gradual care pattern evolution and identifying optimal intervention windows for maximizing sustained ED diversion.

## Data Availability

The data underlying this study cannot be made publicly available as it contains protected health information from the University of Virginia health system and is subject to institutional data use restrictions.

