## Supplemental Table 1 for "Using local and statewide Electronic Health Record data to evaluate the impact of telemedicine in Virginia"

### SUPPLEMENTARY MATERIAL

**Table 5.** Baseline Demographic and Clinical Characteristics of Full Cohort and for both analyses.

| | | Full Cohort | Analysis 1: XGBoost | Analysis 2: Fixed Effects |
|---|---|---|---|---|
| n | | 163247 | 61249 | 59040 |
| sex, n (%) | Female | 97110 (59.5) | 37244 (60.8) | 35984 (60.9) |
| | Male | 66137 (40.5) | 24005 (39.2) | 23056 (39.1) |
| race_ethnicity, n (%) | Asian Non-Hispanic | 2880 (1.8) | 852 (1.4) | 941 (1.6) |
| | Black or African American Non-Hispanic | 19453 (11.9) | 8579 (14.0) | 7976 (13.5) |
| | Hispanic or Latino Any Race | 8977 (5.5) | 3632 (5.9) | 3877 (6.6) |
| | Other Non-Hispanic | 5676 (3.5) | 1840 (3.0) | 2084 (3.5) |
| | Unknown | 2551 (1.6) | 361 (0.6) | 734 (1.2) |
| | White Non-Hispanic | 123710 (75.8) | 45985 (75.1) | 43428 (73.6) |
| Neurodevelopmental_Disorders, n (%) | 1 | 17542 (10.7) | 6444 (10.5) | 6008 (10.2) |
| PVD_or_Hyperlipidemia, n (%) | 1 | 42530 (26.1) | 17871 (29.2) | 14350 (24.3) |
| Seizure_Disorders, n (%) | 1 | 4617 (2.8) | 2041 (3.3) | 1775 (3.0) |
| Chronic_Pain, n (%) | 1 | 42713 (26.2) | 19803 (32.3) | 16265 (27.5) |
| Sleep_Disorders, n (%) | 1 | 38040 (23.3) | 16477 (26.9) | 13704 (23.2) |
| PreDiabetes_Diabetes_or_Obesity, n (%) | 1 | 45354 (27.8) | 19917 (32.5) | 16911 (28.6) |
| ED_visits_before_TM, mean (SD) | | 1.9 (4.0) | 1.9 (3.3) | 1.8 (3.1) |
| ED_visits_after_TM, mean (SD) | | 1.3 (2.6) | 1.2 (2.1) | 1.1 (2.0) |
| pre_index_observation_period, mean (SD) | | 1890.2 (1647.2) | 2263.0 (1683.2) | 1920.7 (1748.0) |
| total_pre_index_visits, mean (SD) | | 32.8 (47.1) | 39.2 (49.8) | 31.6 (46.3) |
| post_index_observation_period, mean (SD) | | 682.3 (655.6) | 594.6 (413.1) | 493.6 (358.9) |
| total_post_index_visits, mean (SD) | | 20.6 (29.5) | 18.6 (24.3) | 17.4 (22.0) |
| CC National Z-Score, mean (SD) | | -0.5 (0.4) | -0.5 (0.4) | -0.5 (0.4) |
| CCI_score_up_through_index_date, mean (SD) | | 1.3 (1.9) | 1.5 (2.0) | 1.3 (1.9) |
| urban_flag, n (%) | 1 | 85022 (52.1) | 31763 (51.9) | 29393 (49.8) |
| if_swva, n (%) | 1 | 5455 (3.3) | 1457 (2.4) | 1805 (3.1) |
| MEDICARE, n (%) | 1 | 27584 (16.9) | 11843 (19.3) | 10611 (18.0) |
| MEDICAID, n (%) | 1 | 20457 (12.5) | 10255 (16.7) | 10046 (17.0) |
| pre_index_ed_duration, mean (SD) | | 544.8 (410.1) | 526.7 (388.2) | 502.9 (349.0) |
| post_index_ed_duration, mean (SD) | | 850.1 (661.8) | 634.3 (421.0) | 572.1 (352.2) |
| vhi_ed_visits_total_pre_index, mean (SD) | | 1.3 (3.5) | 2.4 (4.3) | 2.5 (4.3) |
| vhi_ed_visits_total_post_index, mean (SD) | | 2.2 (5.7) | 2.8 (5.6) | 2.7 (5.0) |
| pre_index_vhi_ed_total, mean (SD) | | 1.3 (3.5) | 2.4 (4.3) | 2.5 (4.3) |
| post_index_vhi_ed_total, mean (SD) | | 2.2 (5.7) | 2.8 (5.6) | 2.7 (5.0) |
| pre_index_ed_visits_ddiff_mean, mean (SD) | | 42.1 (123.5) | 77.1 (154.5) | 79.2 (149.7) |
| age_range, n (%) | 0-17 (Minor) | 24384 (14.9) | 8012 (13.1) | 7727 (13.1) |
| | 18-24 (Young Adult) | 10696 (6.6) | 3964 (6.5) | 4306 (7.3) |
| | 25-34 (Adult) | 18811 (11.5) | 7116 (11.6) | 7591 (12.9) |
| | 35-44 (Middle-Aged Adult) | 19088 (11.7) | 7057 (11.5) | 7218 (12.2) |
| | 45-54 (Senior Adult) | 20197 (12.4) | 7597 (12.4) | 7242 (12.3) |
| | 55-64 (Pre-Retirement) | 24763 (15.2) | 9521 (15.5) | 8696 (14.7) |
| | 65+ (Senior Citizen) | 42693 (26.2) | 17440 (28.5) | 15698 (26.6) |
| | None | 2615 (1.6) | 542 (0.9) | 562 (1.0) |
